# Analysis of Hepatitis B virus genotype D in Greenland suggests presence of a novel subgenotype

**DOI:** 10.1101/2020.06.06.20123968

**Authors:** Adriano de Bernardi Schneider, Reilly Hostager, Henrik Krarup, Malene Børresen, Yasuhito Tanaka, Taylor Morriseau, Carla Osiowy, Joel O. Wertheim

## Abstract

A disproportionate amount of Greenland’s Inuit population is chronically infected with Hepatitis B virus (HBV; 5-10%). HBV genotypes B and D are most prevalent in the circumpolar Arctic. Here, we report 39 novel HBV/D sequences from individuals residing in southwestern Greenland. We performed phylodynamic analyses with ancient HBV DNA calibrators to investigate the origin and relationship of these taxa to other HBV sequences. We inferred a substitution rate of 1.4×10^−5^ [95% HPD 8.8×10^−6^, 2.0×10^−5^] and a time to the most recent common ancestor of 629 CE [95% HPD 37-1138 CE]. The Greenland taxa form a sister clade to HBV/D2 sequences, specifically New Caledonian and Indigenous Taiwanese samples. The Greenland sequences share amino acid signatures with subgenotypes D1 and D2, and approximately 98% sequence identity. Our results suggest the classification of these novel sequences does not fit within the current nomenclature. Thus, we propose these taxa be a novel subgenotype.

## Introduction

Hepatitis B (HBV) is an ancient virus that remains a substantial public health burden. Transmitted by bodily fluid, it is one of the most prevalent viruses in the world, infecting over 2 billion people, 257 million of whom are chronically infected (1). According to the World Health Organization, infection in infancy and early childhood leads to chronic HBV in about 95% of cases, while infection in adulthood leads to less than 5% (1).

HBV is classified into 9 distinct genotypes (A through I) and a putative 10th genotype (J) (2). Precedent sorting of genotype identity is calculated by a distance-based sequence threshold of 8% (3–8). Subgenotypes are assigned based on a 4% threshold across the full genome (9), though new phylogenetic inference methods to determine subgenotype have been suggested (10). McNaughton et al. (9) notes that all genotypes have subgenotype sequence divergence rates of 3-8%, with the exception of genotype D, which is 2-4%, indicating a lack of long evolutionary branches separating well-defined clades.

HBV was considered hyperendemic in the western circumpolar Arctic (Alaska, Canada and Greenland) prior to the introduction of of HBV vaccination programs (11), with the burden of viral hepatitis disproportionately borne by Indigenous populations. Prior to the introduction of HBV vaccination programs in the Canadian Arctic in the 1990s, HBV prevalence was about 3% among the Inuit (12); more, the risk of lifetime exposure to HBV was 25% or 5 times higher than the non-Indigenous population (13). Approximately 5-10% of Greenland’s Inuit population remains chronically infected with HBV (14, 15). Since 1965, serosurveys of HBV exposure among Greenland residents have demonstrated high rates of exposure among adults and chronic infection rates 14-40 times higher than the US and northern Europe (13). Genotypes B and D predominate in the North American circumpolar Arctic (12). Western Greenland’s infections are represented by two genotypes: a majority genotype D and a minority genotype A (12). While infancy is regarded as the most receptive stage of HBV infection, incidence of active infection increases in adolescence (13). This highlights transmission by sexual contact, the primary method for HIV transmission in Greenland (16).

The most prevalent and endemic subgenotype, B5 (formerly B6), within the Alaska Native and Inuit populations of the western circumpolar Arctic may have originated coincident with the rapid movement of the Thule from regions of Alaska to the eastern Arctic (17) approximately 1000 years before present (YBP). Greenland Inuit harboring genotype D HBV (HBV/D) infections, discussed in this study, may represent another endemic lineage with high rates of chronic infection and, importantly, a unique coalescent origin from other defined HBV/D subtypes. Known presence of HBV/D in the Arctic is limited to Greenland (18), Western Canada, and Alaska (19, 20). Association of HBV/D with First Nation Dene living in the western Arctic has been observed, suggesting a separate introduction from Greenlandic Inuit (19). Additionally, Inuit residing in the Canadian Arctic, infected with subgenotype HBV/B5 experience less severe clinical outcomes typical of non-Indigenous people infected with other HBV/B subgenotypes (21). This reduced risk phenomenon for Inuit infected with HBV/D is unknown. Historically, genotype D has poorer clinical prognoses and a lower response to therapy (10, 22). Specific basal core promoter (BCP) nucleotide mutations, T1762 and A1764, are important markers associated with liver disease progression and development of hepatocellular carcinoma (HCC) (23), and are highly associated with HBV genotype D (4).

Despite its broad geographical distribution and large volume of sequence data, the evolutionary history of HBV remains ambiguous. Difference in substitution rates depending on whether the infection is acute, chronic, or within-versus-between hosts (24) is compounded by discrepant calibrations (i.e. fossil-based or tip-dated). Ancient DNA (aDNA) calibration for exogenous viruses has been shown to tease apart some of the evolutionary mystery of HBV (25), revealing a surprising lack of temporal genetic change in the last half millennium (26). HBV is a suitable candidate for aDNA external calibration because of its high viremic levels, even during periods of prolonged infection, and covalently closed circular DNA (cccDNA) genome structure (2).

In this study, we report thirty-nine novel Greenland taxa and investigate their phylogenetic relationships and the time to the most recent common ancestor (TMRCA) of HBV/D in Greenland. To understand how and when HBV reached Inuit populations, we analyzed novel sequence data combined with publicly available datasets and ancestral sequences. Here, we investigate the Greenland clade’s genetic composition and existing genotype D heterogeneity and present the description of a novel subgenotype.

## Methods

### Data collection

Thirty-nine serum specimens were collected from twenty-five individuals infected with HBV from 5 settlements in Western Greenland: Aasiaat, Itilleq, Nuuk, Sarfannguaq and Sisimiut (Figure 1), as previously described (17, 27) (Supplementary Table S1). In certain cases consecutive paired sera separated 5 to 10 years were collected from individuals for the purpose of investigating the intrapatient genetic diversity of HBV (27). All specimens were collected on dates ranging from 1998 to 2017 (Supplementary Table S1). Viral genomic DNA was extracted from 200 µL sera and amplified as previously described (17). Amplicons were sequenced with an AB 3730 XL DNA Analyzer using Big Dye terminator chemistry (Thermo Fisher Scientific, Burlington, ON, Canada). Sequences were assembled and analyzed using DNA sequence analysis software (Lasergene software suite v 15.0.0, DNASTAR, Madison, WI, USA).

**Fig. 1.**
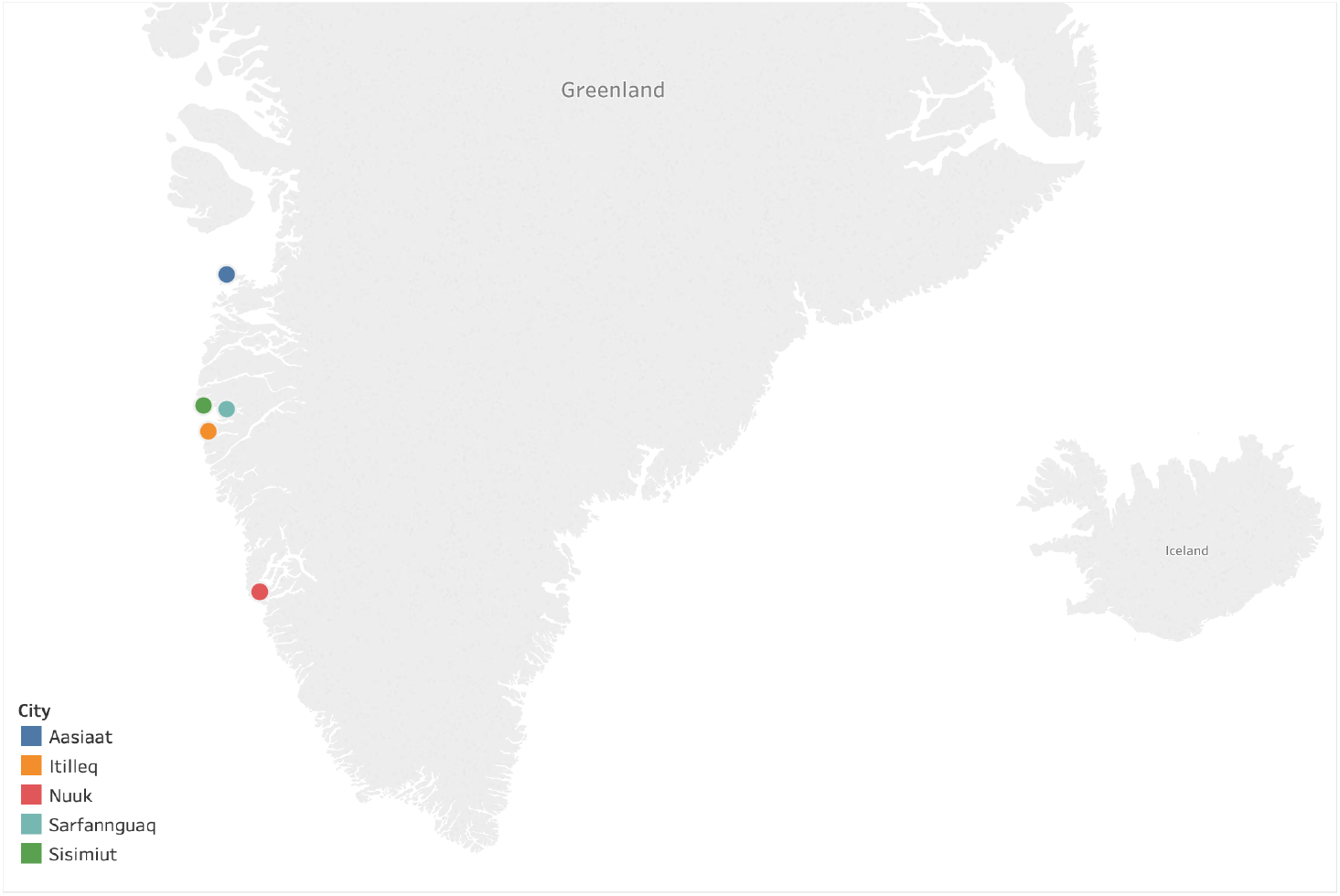
Sampling locations of Hepatitis B virus (HBV) isolates within Western Greenland. The map was generated using Tableau Desktop 2009.1 (www.tableau.com).

Ethical approval for specimen collection was granted by the Statens Serum Institute (Copenhagen, Denmark) and the Commission for Scientific Research in Greenland (Approval number 505-99). Written informed consent was obtained from each participant.

### Taxon sampling

We created two datasets for the analysis of the Greenland taxa. The first dataset was 3260 nucleotides in length and included 54 sequences from all HBV genotypes in order to assess the clustering of the Greenland taxa within existing HBV diversity. Subgenotypes 1 and 2 of genotypes A-D were included, and two genotype E sequences, thereafter a single representative sequence of each genotype F-J. The second dataset was 3182 nucleotides in length and included 93 sequences. We kept two identical sequences which were serially collected from a single Greenlandic patient with Taxon IDs 1205 sampled in 1998, and 312000479 sampled in 2004. Out of the 39 Greenlandic sequences, 29 are novel and are being published here for the first time (Supplementary Table S1). Seventeen sequences cover the full genome, while the remaining twelve are partial or nearly complete, though at least 1253 nucleotides in length. We selected 54 sequences to represent HBV/D history and calibrate our data (Supplementary Table S1). Forty-two sequences serve as global representation of all subgenotypes (D1, n=10; D2, n=11; D3, n=5; D4, n=6; D5, n=5; D6, n=5). An additional four are “novel quasi-subgenotype D2 of hepatitis B virus” full length genome sequences from Taiwanese Indigenous peoples ((28) AB555496, AB555497, AB555500, AB555501), two are from New Caledonia (HQ700511) and Argentina (JN688695) outgroup to the Aboriginal taxa, and six are partial or nearly complete ancient HBV (aHBV) DNA sequences at least 943 nucleotides in length. Four aHBV sequences came from the Mühlemann et al. (25) study (LT992438, LT992439, LT992444, LT992454) and two aHBV came from the Krause-Kyora et al. (26) study, a German medieval tooth and an Italian mummy available at PRJEB24921 of the European Nucleotide Archive. The ages of these ancient sequences ranged from 281 Before Common Era (BCE) to 1569 Common Era (CE).

GenBank accession numbers for novel and previously published HBV genomes are included in Supplementary Table S1.

### Molecular sequence analyses

We computed multiple sequence alignments using MAFFT (29) under default settings. We visualized the alignments and trimmed ragged edges with AliView (30). We screened our datasets for recombination using RDP4 (31) applying six algorithms: Geneconv, Bootscan, MaxChi, Chimaera, SiScan, and 3Seq.

The genotype D dataset nucleotide alignment was translated to specific open reading frames for the polymerase and preS2 coding regions to determine the presence of signature amino acids associated with HBV/D subgenotypes within the Greenland sequences. We evaluated the amino acid residues for positions 39 (Pre-S2), 100 and 128 (POL Spacer) and 126 (RT) for all Greenland sequences and compared the observed residues to the predominant amino acids of subgenotypes D1 and D2, according to Yousif and Kramvis (32). In addition, we mapped three distinct nucleotide mutations G1896A, A1762T, and G1764A across all Greenland taxa as these are important markers associated with disease prognosis.

### Genetic distance

To calculate the genetic distance between the Greenland clade and other subgenotypes we extracted the consensus sequence for the new Greenland taxa using the alignment sequence visualizer of Geneious Prime 2019.2.1 (https://www.geneious.com). We calculated the distance matrix for the 39 sequences within the Greenland clade and the consensus sequence using the dist.alignment function of the ‘seqinr’ package in R (33). The most closely related sequence to the consensus sequence was selected to represent the entire Greenland clade, following McNaughton et al. (9)’s strategy. Pairwise alignments of the Greenland representative sequence with representative sequences for subgenotypes D1 and D2 were compared separately. Resulting genetic distance between the three sequences was used to investigate the feasibility of classifying the Greenland taxa as a novel subgenotype.

### Phylogenetic tree search

To assign the novel HBV sequences from Greenland to the correct genotype we conducted a phylogenetic tree search under the optimality criterion of maximum likelihood with substitution model testing as available in IQ-TREE (34). We executed a 1000-replicate SH-aLRT-like measure of support (35).

We used BEAST v1.10.4 (36) to calculate the TMRCA of the Greenland taxa on the genotype D dataset. Based on the partial fragmentation of our alignment (see supplementary data) we elected to treat the alignment as a single substitution partition, contrary to the eight-partition approach used on genotype B by Bouckaert et al. (17). We selected the GTR+Γ_4_ substitution model with four empirical base frequencies, a Skyride coalescent tree prior (37), and an uncorrelated relaxed lognormal clock (URL). We also tested a strict clock, however we did not observe convergence in Tracer (38), confirming the URL clock as the appropriate selection. We ran the model in duplicate with a Markov chain Monte Carlo length of 100 million. Convergence was determined by Effective Sample Sizes of at least 200 per statistic.

The final maximum clade credibility tree with metadata annotation was rendered using FigTree (39).

All supplementary data can be found at GitHub (https://github.com/abschneider/Paper_HBV_Greenland).

## Results

### Maximum likelihood tree indicates Greenland lineage independence

Model testing prior to IQ-TREE building identified the best fit model from Akaike and Bayesian Information Criterion as GTR+Γ_4_ with empirical base frequencies. The maximum likelihood tree from the first dataset has the Greenland taxa in a monophyletic group sister to genotype D2 sequences with 99.9% SH-aLRT-like support (see supplementary data). Thus, we assigned them to genotype D for our downstream analyses. All subsequent references to our data refer to the second dataset containing only genotype D sequences.

### Recombination

We excluded no taxa based on recombination. RDP identified two potential recombinants (see supplemental files). The first we interpret to be false identification of the recombinant region from nucleotide position 1720-2579 in eleven D4/D6 sequences containing an 859 base pair section from D2 sequence AB267090. Based on the location of the areas internal and external to the recombination site in UPGMA trees, we see no evidence for inter-subgenotype recombination. An additional sequence, JN792912, that was labelled as mostly recombinant with unknown origin, was removed in addition to the eleven sequences and RDP was re-run. The same region was identified in all D3 sequences instead of D4/D6 with a supposed D1 parent. We hypothesize this is an artifact given the high nucleotide conservation of this region. The UPGMA tree topologies were not suspicious of recombinant template-switching in these taxa.

The second recombinant region flagged by RDP was in an Indian D5 sequence, GQ205382, with a 269 base pair region of a D3 sequence (KX827292) from nucleotides 1765-2034. This region has no mutations between the two sequences. Instead, the D3-specific single nucleotide polymorphisms (SNPs) are found in this D5 sequence, potentially representing a recombination event that cannot be excluded. However, the D5 sequence still clusters with other Indian D5 sequences in a clade with 100% bootstrap in the ML tree. Neither of these taxa produced interference on ML tree topology. Thus, we elected to keep all of the 93 taxa. We found no recombination between D1, D2 and the Greenland sequences.

### Amino acid signatures

The Greenland HBV/D amino acid signatures on Pre-S2 aa39, POL Spacer aa100 and aa128, and RT aa126 introduces a paradigm in relation to D1 and D2 signatures (Table 1). The Greenland sequences do not “fit” into either D1 or D2 subgenotypes amino acid signatures predominantly observed with these subgenotype sequences according to Yousif and Kramvis (32). Instead, they share characteristics of D2 at aa39 and aa100, and D1 at aa126 and aa128. As the Greenland sequences are monophyletic, this suggests the presence of a lineage with a new amino acid signature.

**Table 1.** Amino acid signatures of HBV genotype D1, D2 and Greenland taxa. Adapted from (32), Table 2.

| AA Signature | D1 | D2 | Greenland |
| --- | --- | --- | --- |
| Pre-S2 aa39 | V/a | A/v | A |
| POL Spacer aa100 | S/a, t | A/t, v | A |
| POL Spacer aa128 | S/g | G/s | S |
| RT aa126 | H | R/h | H |

### Mutations

We observed mutations in our sequences at positions 1762 (wild type A; mutant T), 1764 (wild type G; mutant A) and 1896 (wild type G; mutant A). The 1762 and 1764 site mutations are associated with a significant increased risk of liver disease progression and hepatocellular carcinoma development (23, 40). The mutation on site 1896 is associated with HBeAg negativity and seroconversion to anti-HBeAg positivity by introducing a stop codon to the HBeAg reading frame (41). Twenty two of the 39 Greenlandic sequences had a G to A mutation at nucleotide position 1896. Of the 13 consecutively sampled taxa, only two pairs reflected this substitution, whereas most (n=5) had a mutant A at position 1896 in both samples. Mutations A1762T and G1764A were not as prevalent, present in only 9/39 and 14/39 Greenlandic sequences, respectively (Table 2).

**Table 2.**
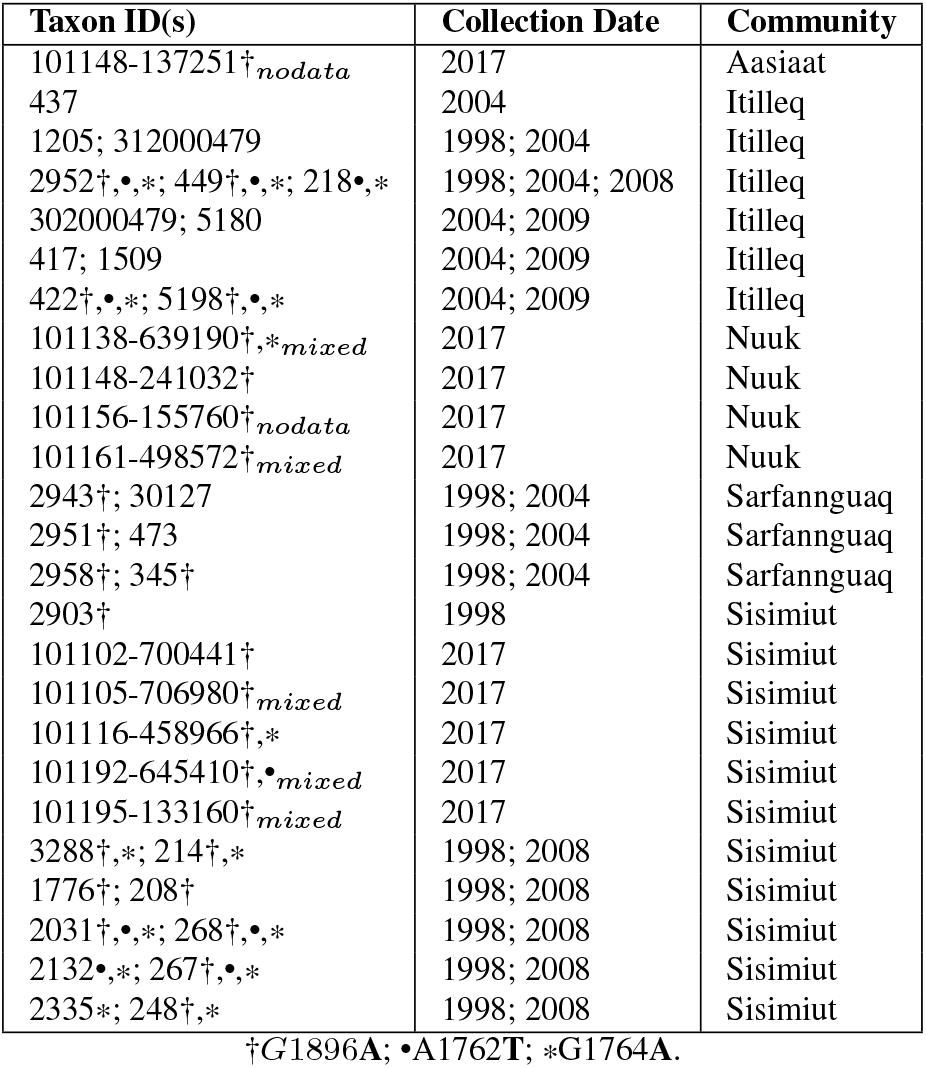
Greenland HBV/D taxa and consecutive paired sera indicated by multiple taxon IDs in a single cell separated by semicolon. Significant mutant nucleotides (bolded) are indicated by symbol, opposed to unmarked wild type nucleotides.

### Genetic distance

Our consensus sequence to the Greenland clade had 100% identity to six sequences: 101102-700441, 417, 302000479, 5180, 1205, and 312000479. We utilized the same sequences as McNaughton et al. (9) as references for the comparison of subgenotypes: One from each of the two major D1 clades: AB222711.1 (1), KC875277.1(2) and a single D2 sequence: MF925358.1. The pairwise alignments between the reference sequences and the Greenland sequences resulted in a pairwise identity of 98.1% for D1(1), 98.3% for D1(2) and 97.9% for D2 (Table 3).

**Table 3.** Genetic distance between our reference Greenland sequence and representative taxa of D1 (two major clades) and D2 subgenotypes.

| GL taxon | Reference sequence | Seq Identity | Divergence |
| --- | --- | --- | --- |
| 1205 | AB222711 (D1) Clade 1 | 98.1% | 1.9% |
| 1205 | KC875277 (D1) Clade 2 | 98.3% | 1.7% |
| 1205 | MF925358 (D2) | 97.9% | 2.1% |

### Substitution rate and TMRCA of HBV/D in Greenland populations

We inferred on the molecular clock analysis a substitution rate of 1.398 × 10 ^−5^ [95% HPD 8.837 × 10 ^−6^, 1.952 × 10 ^−5^], similar to that obtained by Mühlemann et al. (25): 1.18 × 10 ^−5^ [9.21 × 10 ^−6^,-1.45 × 10 ^−5^]. The age of the root, representing genotype D, was 675 BCE (1221 BCE, 281 BCE). The age of the Greenland taxa was 629 CE (95% HPD interval 37 CE, 1138 CE).

## Discussion

Studies on HBV phylogenetics provide a means of understanding the history of the disease spread in an epidemiological context. In this study, we reveal a novel subgenotype of HBV genotype D isolated from Greenland. Interestingly, this novel clade sister to subgenotype D2 shares genetic characteristics of both subgenotypes D1 and D2. Inference of all Greenland taxa as monophyletic in maximum likelihood and Bayesian frameworks supports the hypothesis of a single origin event giving rise to a distinct and isolated lineage on the west coast of Greenland.

Phylogeography in the Greenland dataset was not used given the limited geographic scope of the Greenland taxa and lack of logistical purpose exploring global movement given our reductive representation of genotype D taxa. Twenty two of the Greenlandic sequences shared a G to A substitution at nucleotide position 1896, associated with seroconversion of positive HBeAg reactivity to anti-HBeAg positivity, and historically most present in genotype D (42). These taxa likely had higher rates of evolution (41), though our methods were unable to tease apart their contribution to the substitution rate. Of our consecutively sampled pairs/triplet, the lack of pre-core mutation (n=3) or a substitution from the mutant to the wild type (n=3) indicates active infection occurring over multiple years, though our lack of sampling only offers sporadic glances into infection dynamics. The identical pair of consecutively sampled sera separated by six years is interesting, as we expected this individual to seroconvert over this time span. These findings reflect the slow and tangled nature of HBV evolution.

For instance, though no recombination was inferred between the Greenland sequences and subgenotype D1/D2, the over-lapping serotype signatures (Table 1), suggest that the Green-land taxa cannot be neatly fit into the amino acid composition of either subgenotype.

HBV subgenotypes within genotype D are not well defined by the precedent method of genetic distance, as is the case for most HBV classification (9). It has become increasingly apparent that nucleotide divergence interferes with phylogenetic interpretation (10). The Greenland taxa’s inferred sister clade, subgenotype D2, suggests a single origin between the Greenland and D2 taxa that is not inclusive of D1, with 94% posterior support (Figure 2).

**Fig. 2.**
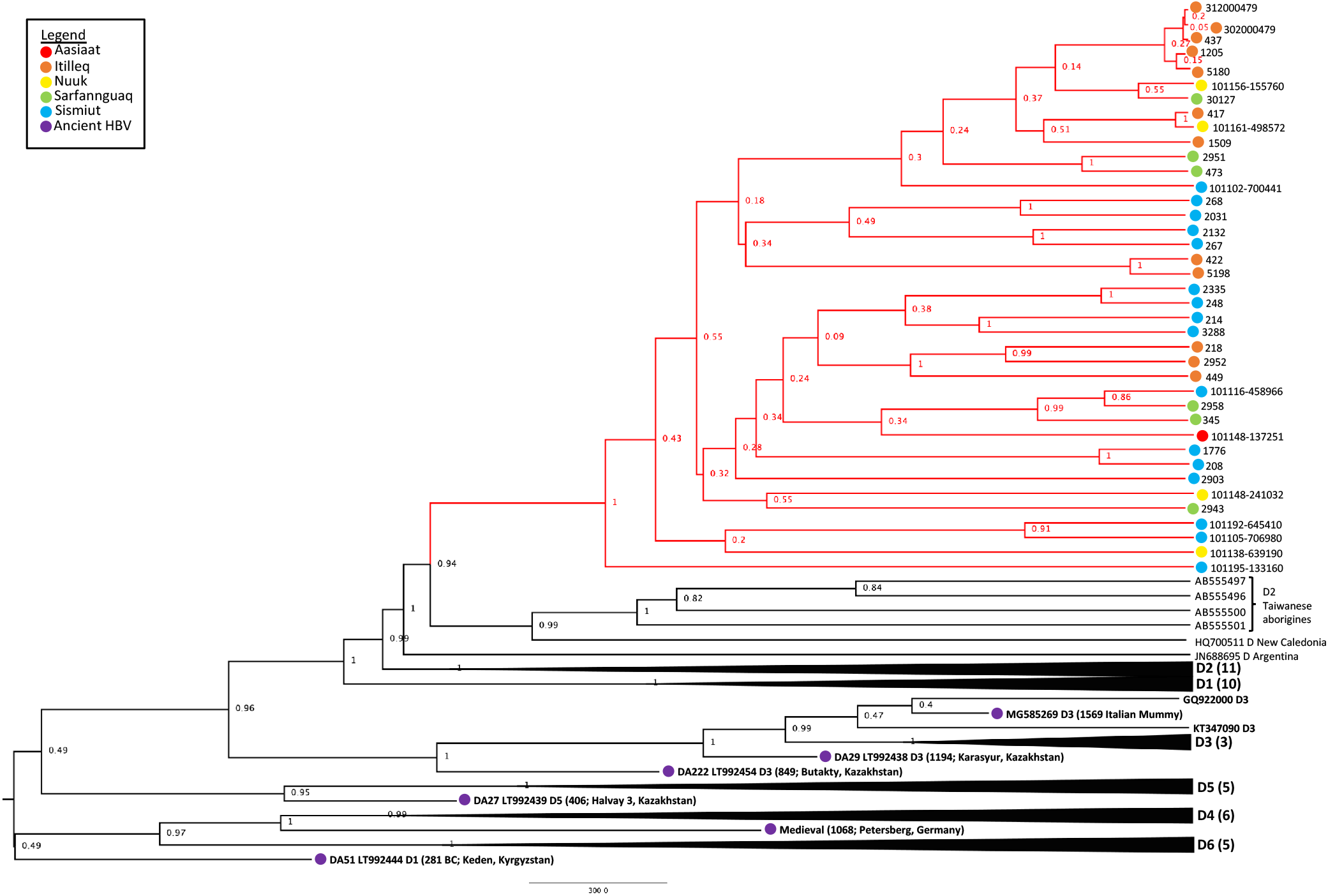
HBV/D Maximum Clade Credibility (MCC) tree from combined run trees with posterior support indicated on nodes. Greenland lineages are shaded in red and corresponding location indicated by colored circle on branch tips.

Taxa are likely to be geographically related, preserving the complication that may be faced with pure geographic assignment (43, 44). Rather than focusing on endemicity, location, or solely genetic distance, the best way to characterize HBV may be by observing evolutionarily independent lineages through phylogenetic analysis, as is argued by Peterson (45). Phylogenetic analysis circumvents the use of genetic distance as a sole metric for sub-/geno-type naming. Further, while genetic distances between the Greenland taxa and subgenotypes D1 and D2 (1.7-1.9% and 2.1%, respectively) are on the threshold of what McNaughton et al. (9) cites as the majority of genotype D pairwise distance (i.e. 2-4%, compared to the typical subgenotype divergence of 3-8%), they are distinctly monophyletic across our methods. Here, we present our inference of 39 Greenland HBV/D taxa as a newly characterized subgenotype exemplified by this topologically independent lineage.

The Greenland taxa’s TMRCA of 629.42 CE translates to a coalescence point about 1390 years before present (YBP), preceding the only other Greenland HBV lineage (B5) studied (17). Their study estimated that B5 was introduced to Greenland during coastal route movement of the Thule in the last 1000 years (17), calibrated by the date associated with Thule migration to the Eastern Arctic (647 to 953 YBP, or 1370 to 1064 CE). Nonetheless, they are distinct genotypes evolving at different rates, thus it is feasible that these genotypes arrived in Western Greenland following separate introductions.

The more closely related subgenotype D2 may have an origin from the Middle East (46) or Southern Europe (47). Use of ancient HBV DNA allowed us to estimate a Greenland HBV/D TMRCA range of 37-1138 CE, suggesting the virus was introduced at some point to the Thule, but this ancestor no longer exists. However, ancestral remnants aside from the Taiwanese Indigenous and New Caledonian samples which formed an adjacent clade to the Greenland taxa are necessary for improved confidence in inferring the route which brought HBV/D in these populations.

Integrating novel sequences with published HBV/D data in this study has demonstrated a strongly inferred and geographically independent monophyletic lineage from existing HBV/D subgenotype architecture. We reveal that HBV/D samples from 25 Western Greenland residents, 13 of which were consecutively sampled over 5-10 years, form an evolutionarily unique clade distinct from subgenotypes D1 and D2.

Pairing these modern data with ancient DNA, we uncovered more detail in the story of HBV genotype D evolution. Such diachronic studies are necessary for the tough-to-interpret HBV (25), where study results on rate and origin vary by orders of magnitude subjective to the data and model limitations. Additional HBV/D sequences from ancient archeological remains in the Arctic are necessary to resolve the mystery of its origin and pattern of dispersal beyond speculation.

## Data Availability

All supplementary data can be found at GitHub (https://github.com/abschneider/Paper\_HBV\_Greenland).

## Acknowledgements

The authors acknowledge and are grateful for the technical assistance of Ms. Elizabeth Giles.

